# Changes in back pain scores after bariatric surgery in obese patients: A systematic review and meta-analysis

**DOI:** 10.1101/2020.08.25.20182022

**Authors:** Froukje W. Koremans, Xiaolong Chen, Abhirup Das, Ashish D. Diwan

**Author notes:** **Corresponding author:** Abhirup Das, PhD, Level 3, WR Pitney Building, St. George & Sutherland Clinical School, The University of New South Wales, Kogarah, Sydney, NSW 2217.

## Abstract

**Objective:** To evaluate if back pain scores in morbidly obese patients change after bariatric surgery.

**Summary Background Data:** Obese patients often complain of low back pain (LBP), however the underlying mechanism is not fully understood. Recent research shows that, next to mechanical loading, the chronic low-grade inflammation that arises in obese patients is contributing to LBP due to intervertebral disc degeneration. Therefore, it is hypothesized that bariatric surgery will have an effect on the LBP in obese patients.

**Methods:** We searched four online databases for randomized controlled trials and observational studies. In obese patients, eligible for bariatric surgery, the changes in pre- and postoperative pain scores, assessed by Numeric Rating Pain Scale (NPS) or Visual Analogue Scale (VAS), were considered as primary outcomes. Effect size (ES) and their 95% confidence intervals (CI) were evaluated.

**Results:** Eight observational studies met the eligibility criteria. All studies showed a reduction of LBP following bariatric surgery, with a mean change of −2.9 points in NPS and of −3.8 cm in VAS. Among the patients undergoing bariatric surgery, based on a fixed effect estimated by pain assessment, the pain score decreased significantly in both groups; in NPS (ES −3.49, 95%CI [−3.86, −3.12]) and in VAS (ES −3.975, 95%CI [−4.45, −3.50]).

**Conclusions:** From this meta-analysis, the data of back pain improvement following bariatric surgery seems encouraging. Substantial weight loss following bariatric surgery might be associated with a reduction in back pain intensity.

## INTRODUCTION

Obesity is a growing health problem worldwide. The WHO reported that about 1.9 billion adults were overweight and 600 million adults were obese in 2016, the numbers which continue to rise over the years.^1^ Obesity is defined as an excessive or abnormal fat accumulation and is calculated by the body mass index (BMI). It is a complex and multifactorial condition, with a great risk for the patients’ health. A BMI above 25 mg/kg^2^ is defined as overweight and a BMI of 30 or more as obese. Obesity is associated with an increased risk for diabetes mellitus type 2, hypertension, cardiovascular diseases, cancer and musculoskeletal diseases, including low back pain (LBP).^2-4^ Among the obese patients the prevalence of LBP ranges from 22% to 68.1%.^5 6^ According to the American Obesity Association (AOA) backpain is prevalent in nearly one-third of the classified obese Americans. Similar to obesity, LBP is a major pressing health problem worldwide.^7^ During their lifetime approximately 80% of the adults experience at least one episode of LBP.^8^ Patients with symptomatic LBP often report major disability and a decrease in quality of life. LBP imposes a high individual and social economic burden that is similar to or even greater than that of other major health problems, such as coronary heart or Alzheimer’s diseases.^9-11^

An important cause of LBP is intervertebral disc degeneration (IDD). IDD has traditionally been considered as an age-related process of decreasing proteoglycan content.^9, 10, 12^ However, recent studies suggest additional contributing factors, such as genetic factors, injuries and smoking, that can expedite IDD.^9, 10, 13, 14^ Another major contributing factor is obesity.^15-18^ A large MRI study showed an intricate relationship between obesity and disc degeneration.^16^ Similar studies showed that a greater fat mass is associated with increased back pain and reduced disc height.^17, 18^ Interestingly, obesity was previously thought to be a mechanical risk factor for causing backpain, but recent evidence suggests that the inflammatory factors of obesity also contribute to LBP in patients.^19-22^ In the last two decennia it has become more evident that many of the comorbidities that arise in obese patients involve a chronic low-grade inflammation. Adipose tissue is recognized as an active endocrine organ, secreting cytokine-like hormones. It is hypothesized that these biochemical factor(s) in obese patients act as mediators for IDD too and this might explain the negative effect of obesity on the homeostasis of the intervertebral discs.^23-25^

The current management of symptomatic IDD is conservative or surgical. Conservative interventions are preferred, but surgical procedures will be chosen for patients who are nonresponsive to conservative interventions or had a progressive neurological impairment or both. Spinal surgery is highly invasive with a great risk of relapse, loss of mechanical properties and degeneration of adjacent segments. On top that, there is no guarantee of resolution of symptoms after surgery.^26^ Beside this, obese patients have a greater pre- and postoperative risks. The complication rate for obese patients is estimated to be 2.5 times greater, mostly due to a longer required surgical time.^27-29^ A positive correlation has been observed between obesity rate and complication rates.^30^ Furthermore, obese patients have a ten-time greater risk for wound complications postoperatively. Considering all of the above, spine surgeons often recommend morbidly obese patients to lose weight prior to elective spine surgery. The majority of those patients eventually seek specialized medical help for weight reduction, which often results in bariatric surgery. Normally, one year after bariatric surgery patients show a total weight loss around 18-30%, depending on the bariatric procedure.^31^ Besides weight reduction, bariatric surgery decreases inflammatory responses and reduces the obesity-associated comorbidities, such as diabetes mellitus type II and the patients increased cardiovascular risk.^32, 33^ Because chronic low-grade inflammation in obese patients contribute to the IDD and LBP, we were wondering if bariatric surgery might also have an influence on the back pain of obese patients. Secondly, we were wondering whether bariatric surgery should be considered prior to spinal surgery in obese patients with non-specific back pain. This meta-analysis aimed to address if back pain scores in morbidly obese patients change after bariatric surgery.

## METHODS

### Search strategies

The literature was searched in accordance with preferred reporting information for systematic reviews and meta-analyses (PRISMA) guidelines.^34^ In November to December 2019 the online databases EMBASE, MEDLINE, PubMed and Cochrane Central Register of Controlled Trials were searched to identify all relevant studies published in English between 1966 and December 2019. The search included the following terms: “bariatrics”, “obesity” “” ‘gastric bypass’, ‘gastric sleeve’, ‘Roux-en-Y’, ‘RYGB’, ‘sleeve gastrectomy’ and ‘adjustable gastric band’ and ‘back pain’, with appropriate combinations of operators “AND”, “OR”, and “NOT” as described in the Supplemental Digital Content 1. Additional references were assessed using the reference lists of relevant studies. The review protocols are registered on PROSPERO (International Prospective Register of Systematic Reviews number, registration number: pending).

### Inclusion criteria

The included studies were:

1. Randomized controlled trials (RCT) and observational studies.
2. Enrolled adult obese patients with a BMI > 40 kg/m^2^, or a BMI ≥ 35kg/m^2^ with comorbidities, undergoing primary bariatric surgery.
3. Assessed the pain intensity change of the low back with Numeric Rating Pain Scale (NPS) or Visual Analogue Scale (VAS) before and after bariatric surgery.
4. Conducted a follow-up of at least 3 months.

### Exclusion criteria

Meta-analysis, systematic review, editorials, *in vitro* biomechanical studies and studies looking into LBP caused by pathological entities were excluded.

### Types of outcomes measures

The following outcomes measures were assessed in this review:

Primary outcome: The change in pain intensity score of the LBP, before and after bariatric surgery, measured in VAS or NPS.

Secondary outcomes: Radiology (disc space height), back-specific disability questionnaires (Rolland-Morris score, Oswestry Disability Index (ODI) and Waddell Disability Index and pain pressure threshold (PPT).

### Selection of studies

Two reviewers (FWK and XLC) screened the titles and abstracts following the inclusion and exclusion criteria. Of all the potential eligible studies the full text was reviewed. All the remaining potential eligible studies were discussed with a third researcher (AD). Disagreements were resolved by consensus. If no consensus was reached the collective advise of our research team was the overriding factor.

### Data collection

Each article was independently abstracted by one reviewer. An overview of the study characteristics was systematically provided. The included data were preoperative and postoperative pain intensity scores, study and patients’ characteristics, type of intervention, pre- and postoperative BMI, follow-up time and the secondary outcomes.

### Risk bias assessment

All the included articles were independently assessed for risk of bias using the Oxford Centre for Evidence-Based Medicine tools (CEBM).^35^ Using the CEBM-tool a level was allocated to each of the eligible studies. Depending on study characteristics this level could range from 1 to 5.

To assess the methodological quality of the included observational studies the Newcastle-Ottawa Scale (NOS) was used.^36^ The NOS-tool judges an article on three domains: (1) selection of the group, (2) comparability and (3) assessment of the outcomes. The score of a study can range from 0-9, where a study with score of 7 or higher will be categorized as low risk. The assessments were discussed with a second researcher, disagreements were resolved by the third reviewer (AD).

### Data synthesis and analysis

We performed two meta-analyses (with and without a fixed effect estimate) to examine the changes in back pain score following bariatric surgery. In this meta-analysis the outcomes were sorted by NPS and VAS scores.

Mean differences and standard errors were calculated to compensate for the great amount of prospective studies without control groups. The mean differences were calculated by subtracting the mean postoperative pain intensity score from the mean pre-operative pain intensity score. Since there is a decrease, the changes were made negative. The standard errors were generated by dividing the standard deviations by the square root of the study population. For the standard deviation the following equation was used:

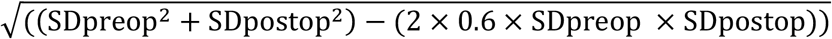

For this meta-analysis STATA software (Release 16, StataCorp LLC, TX) was used for the statistical analyses. Chi-squared (*I*^2^) statistic was used to measure heterogeneity among the trials. *I^2^* < 50% implied homogeneity and the analysis included a random-effects model by the DerSimonian-Laird method. *I^2^* > 50% indicated heterogeneity and, consequently, a fixed-effects model was used according to the Mantel-Haenszel method. We conducted subgroup analysis and sensitivity analysis to assess the impact of heterogeneity. Mean difference and 95% confidence intervals (CI) were reported. A forest plot was used to calculate the effect size (ES). Publication bias was assessed by funnel plot symmetry using the Begg-Mazumdar test. The statistical significance was set at 5% (α = 0.05).

Finally, to calculate the association of mean pain change with BMI change, we included BMI change as a predictor in an meta regression. For this meta-regression proportion of between-study variance was explained with Knapp-Hartung modification.

### Quality assessment

The Grading of Recommendations Assessment, Development and Evaluation (GRADE)- system was used to evaluate the levels of evidence, the quality of assessment and the results from data extraction.^37^ The quality was rated as “very low”, “low”, “moderate” or “high” (Supplemental Digital Content 2).

## RESULTS

The literature search is illustrated in the PRISMA flow diagram (Fig. 1). A total of eight studies that assessed the effect of bariatric surgery on the intensity of back pain in obese patients, measured in NPS or VAS, were included.

**Fig. 1.**
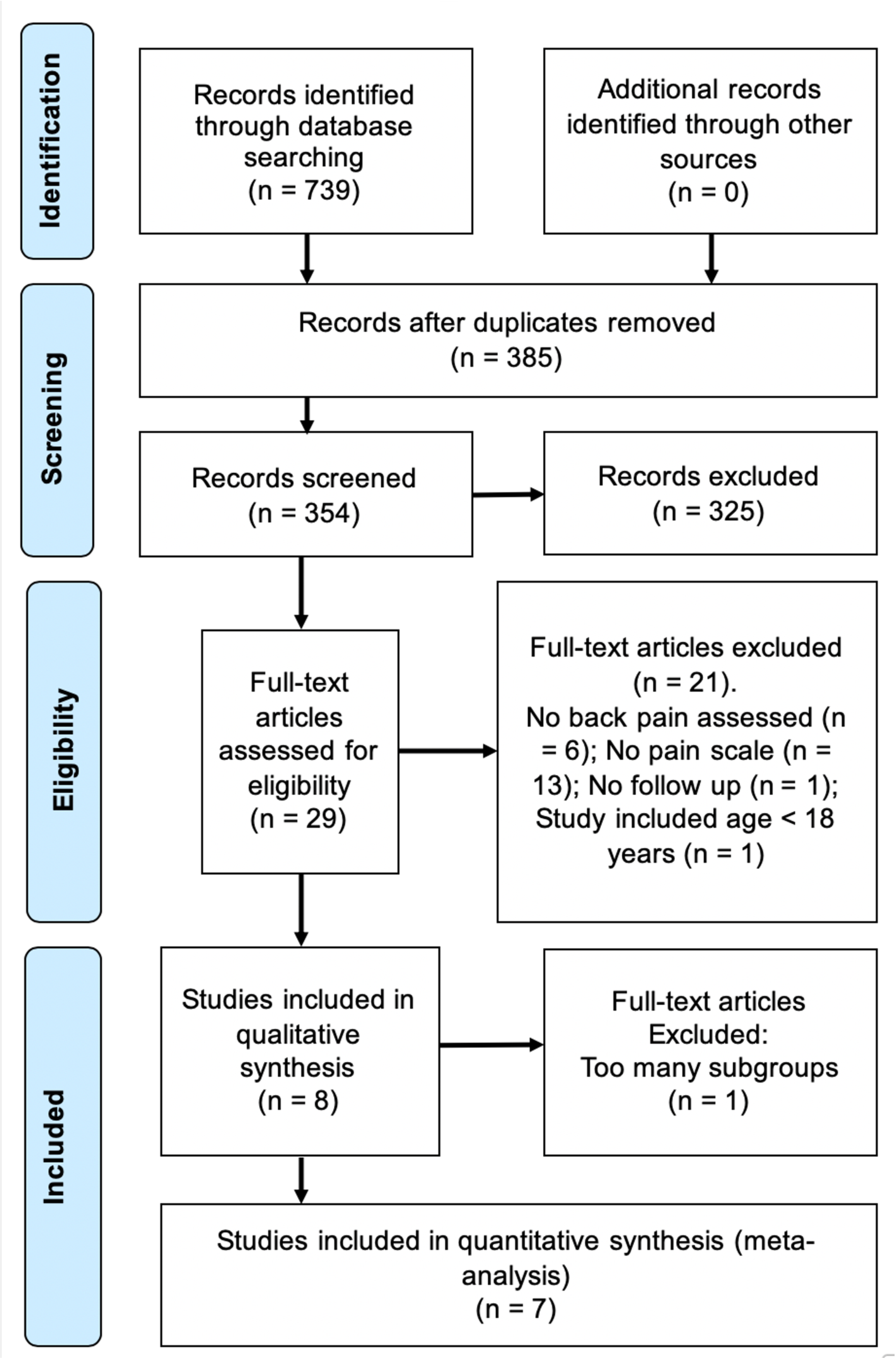
Flow chart showing the selection of articles for back pain evaluation after bariatric surgery in accordance with the Preferred Reporting Items for Systematic Reviews and Meta-analyses (PRISMA) guidelines

### Study characteristics

Among a total of 318 patients in the eligible studies, the median of the mean age was 44.1 years, of which 76.4% were female. Sample sizes ranged from 20 to 72 patients and follow-up times from 3 months to 2 years.

All the eight studies were observational studies, including 3 cohort studies^38-40^ and 5 case series.^41-45^ One progressive study recruited a non-randomized control group,^40^ and seven had no control groups. The bariatric interventions in these studies included Roux-en-Y gastric bypass, sleeve gastrectomy, gastric banding and duodenal switch. Most surgical interventions were performed laparoscopically, some open. Two studies recruited patients with a history of pre-existing back pain,^39, 45^ and six recruited patients with or without back pain.^38, 40-44^ The study characteristics of all included studies are presented in Table 1.

**Table 1.**
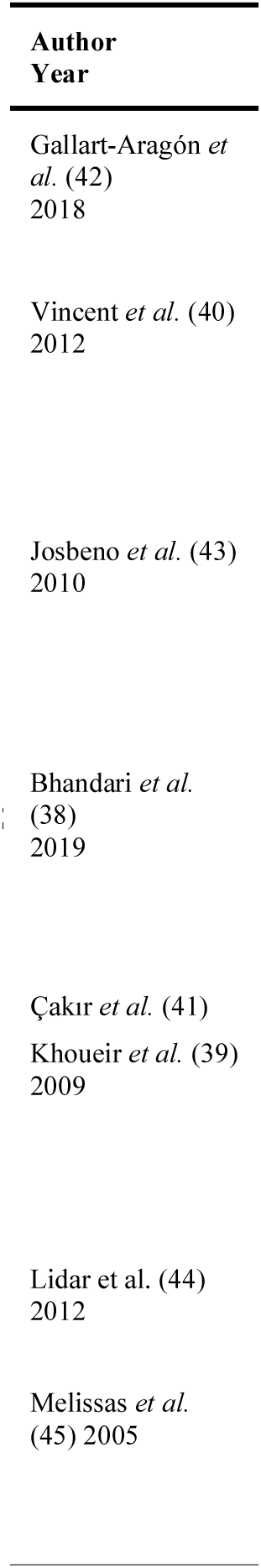
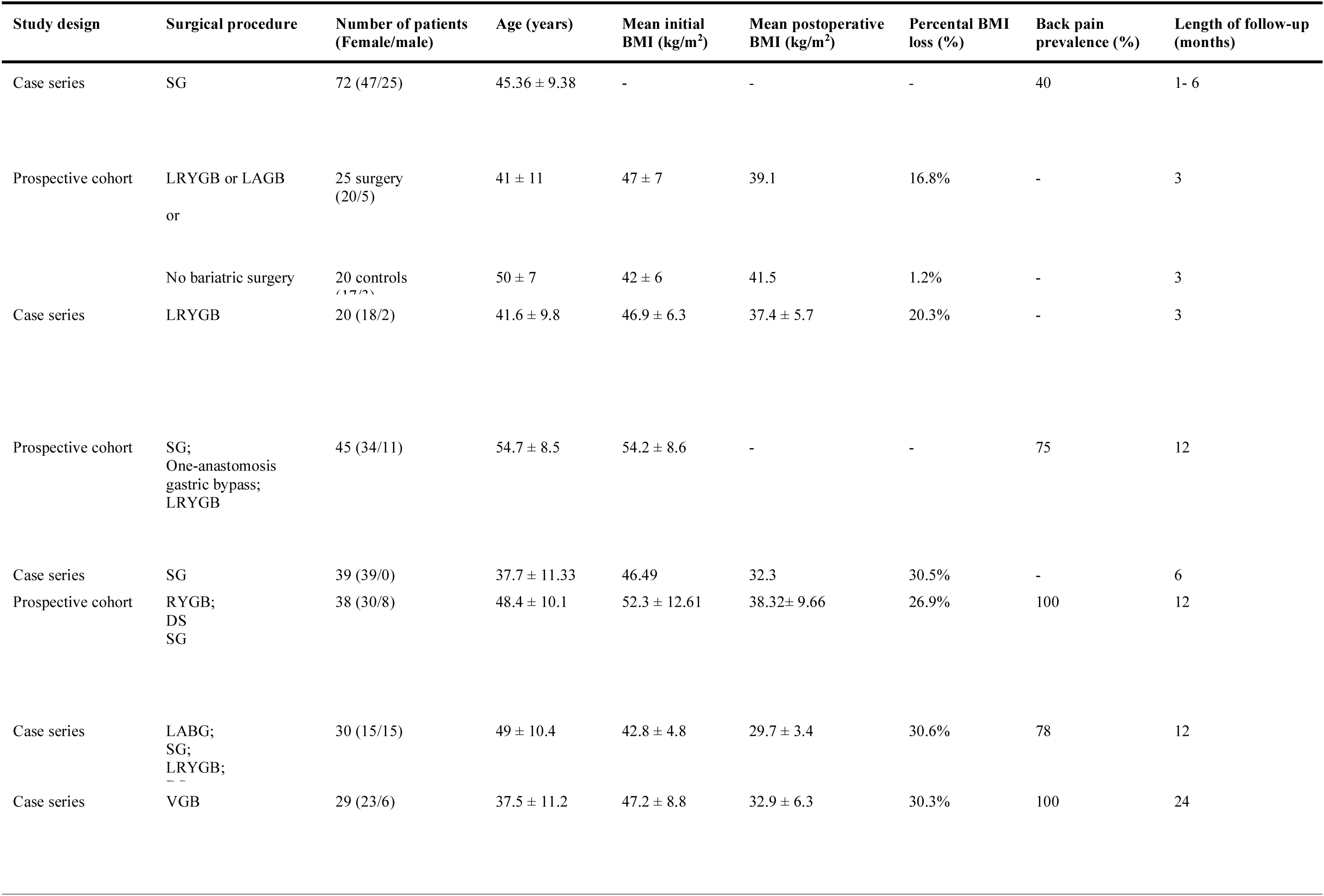
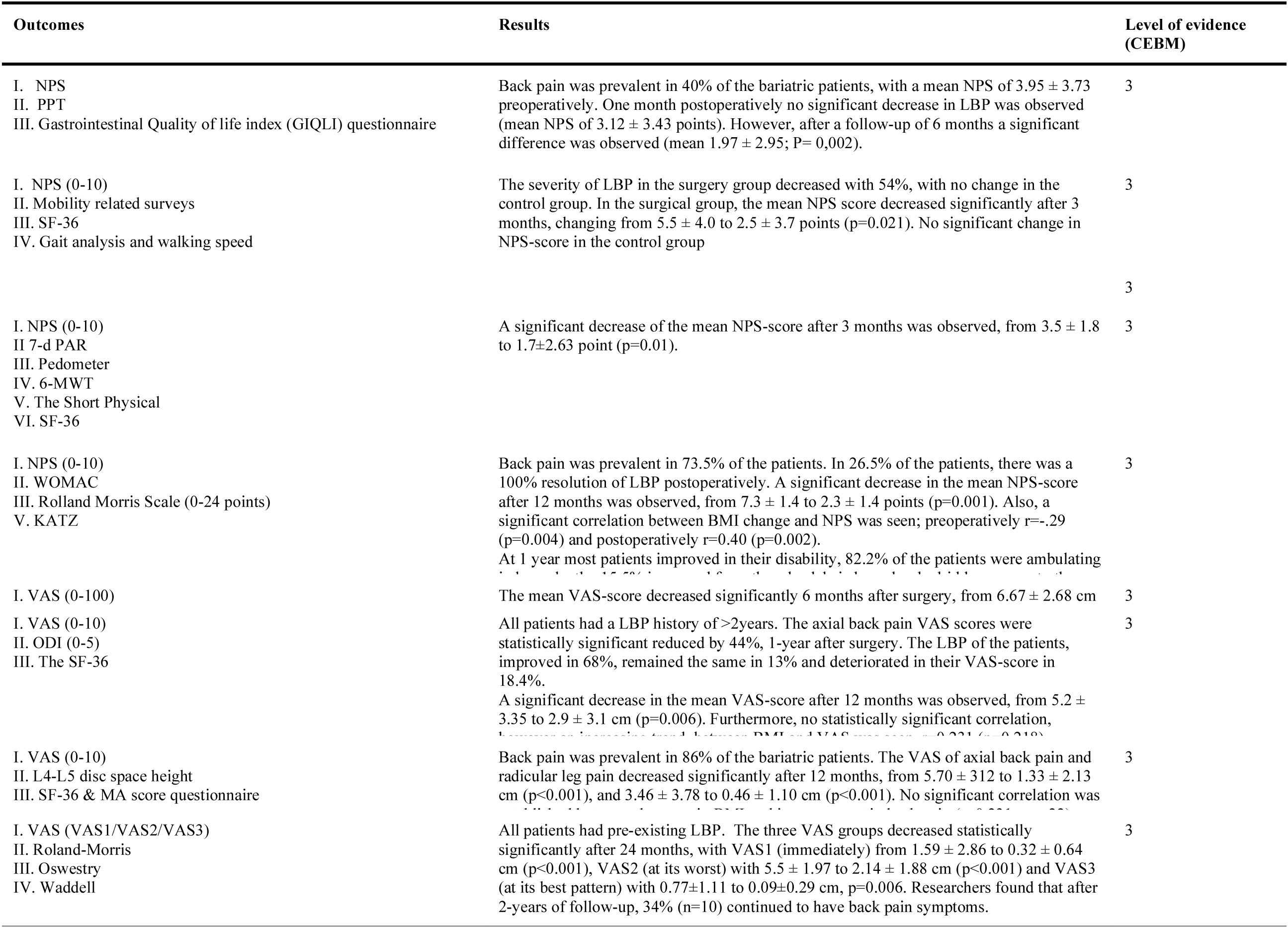
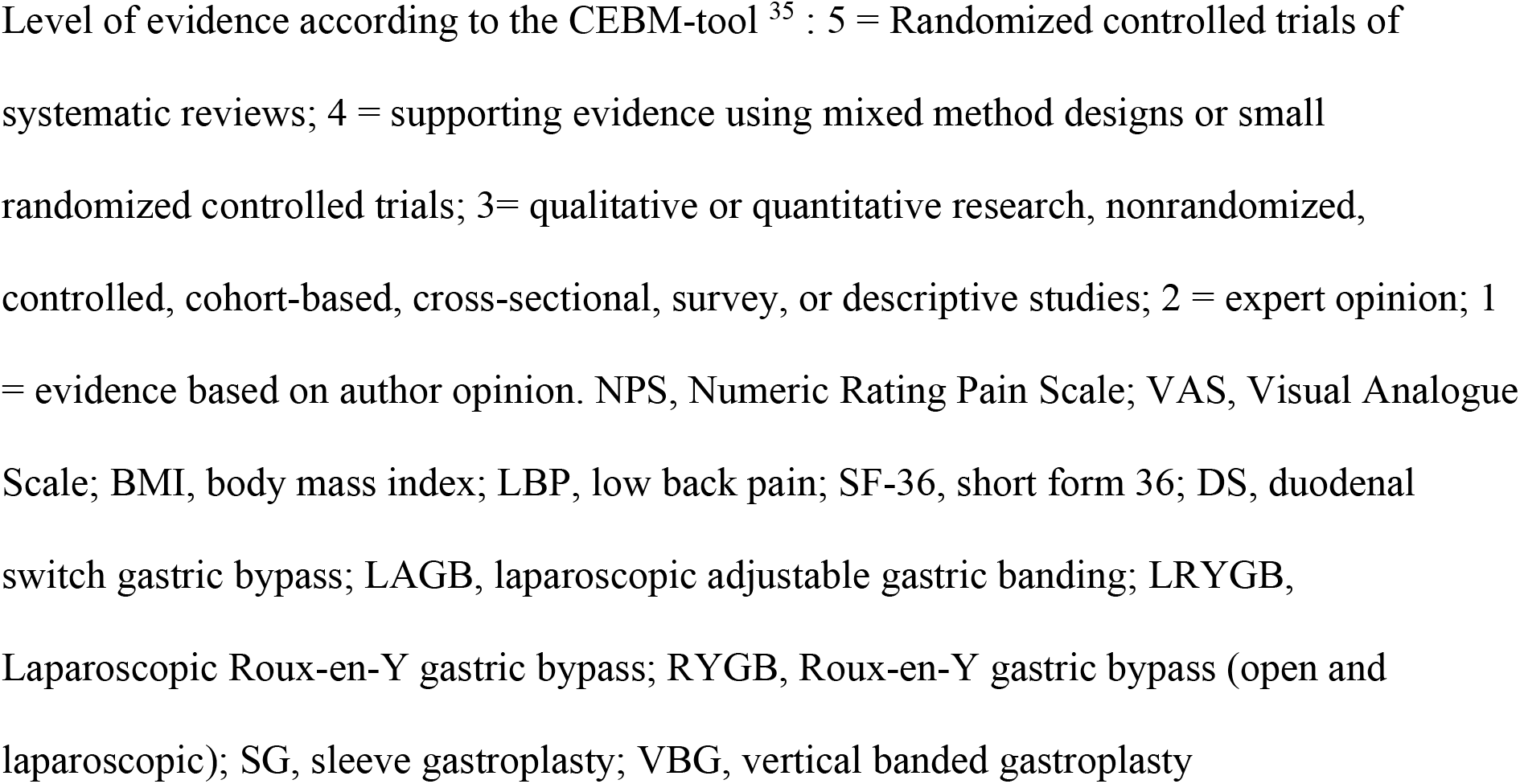
Demographics and results of included studies.

### Risk of bias

According the CEBM-tool all studies were classified as a level 3 study. The results of the bias assessment are shown in Table 1. The methodological quality of the cohort studies was further assessed using the NOS-tools in Table 2. According the NOS only one cohort study was categorized as low risk, with a score of 7, while the other seven observational studies were raided with scores less than seven.

**Table 2.** Assessment of the methodological quality of the studies according to the Newcastle– Ottawa Scale (NOS).^36^

| Author | Year | Country | Surgical procedures | Selection (/4) | Comparability (/2) | Outcome (/3) | Total (/9) |
| --- | --- | --- | --- | --- | --- | --- | --- |
| Gallart-Aragón | 2018 | Spain | SG | 3 | 0 | 2 | 5 |
| Vincent | 2012 | USA | LRYGB or LAGB | 4 | 1 | 2 | 7 |
| Josbeno | 2010 | USA | LRYGB | 3 | 0 | 2 | 5 |
| Bhandari | 2019 | India | SG or one-anastomosis gastric bypass or LRYGB | 2 | 0 | 3 | 5 |
| Çakır | 2015 | Turkey | SG | 2 | 0 | 3 | 5 |
| Khoueir | 2009 | USA | RYGB or DS or SG | 3 | 0 | 3 | 6 |
| Lidar | 2012 | Israel | LAGB or SG or LRYGB or DS | 3 | 0 | 3 | 6 |
| Melissas | 2005 | Greece | VBG | 3 | 0 | 3 | 6 |

A study awarded seven or more stars was regarded as a high-quality study.
BMI, body mass index; DS, duodenal switch gastric bypass; LAGB, laparoscopic adjustable gastric banding; LRYGB, Laparoscopic Roux-en-Y gastric bypass; RYGB, Roux-en-Y gastric bypass (open and laparoscopic); SG, sleeve gastropasty; VBG, vertical banded gastropasty

### Outcomes for Back Pain Intensity

All the studies show a favourable improvement of back pain intensity scores, NPS and VAS, after bariatric surgery. Three uncontrolled studies, without pre-existing back pain in their inclusion criteria, showed that back pain was prevalent in 40 to 86.7% of the patients.^38, 42, 44^ Four studies ^38, 41, 44, 45^ showed that the pain intensity scores were reduced by 70% and the other four studies ^39, 40, 42, 43^ around 50%. Individual results are presented in Table 1. A funnel plot of the results of these 7 trials appeared to be asymmetrical (see Supplemental Digital Content 3_Figure 1, no publication bias).

### Random effect model by Test

Four studies measured pain intensity using NPS.^38, 40, 42, 43^ Among all patients (with or without back pain), the mean back pain intensity score was 2.9 points lower after bariatric surgery compared to before. In our meta-analysis the change in NPS-score after bariatric surgery showed a significant change (ES −2.96, 95%CI [−4.78, −1.15], p=0.000) (Fig. 2).

**Fig. 2.**
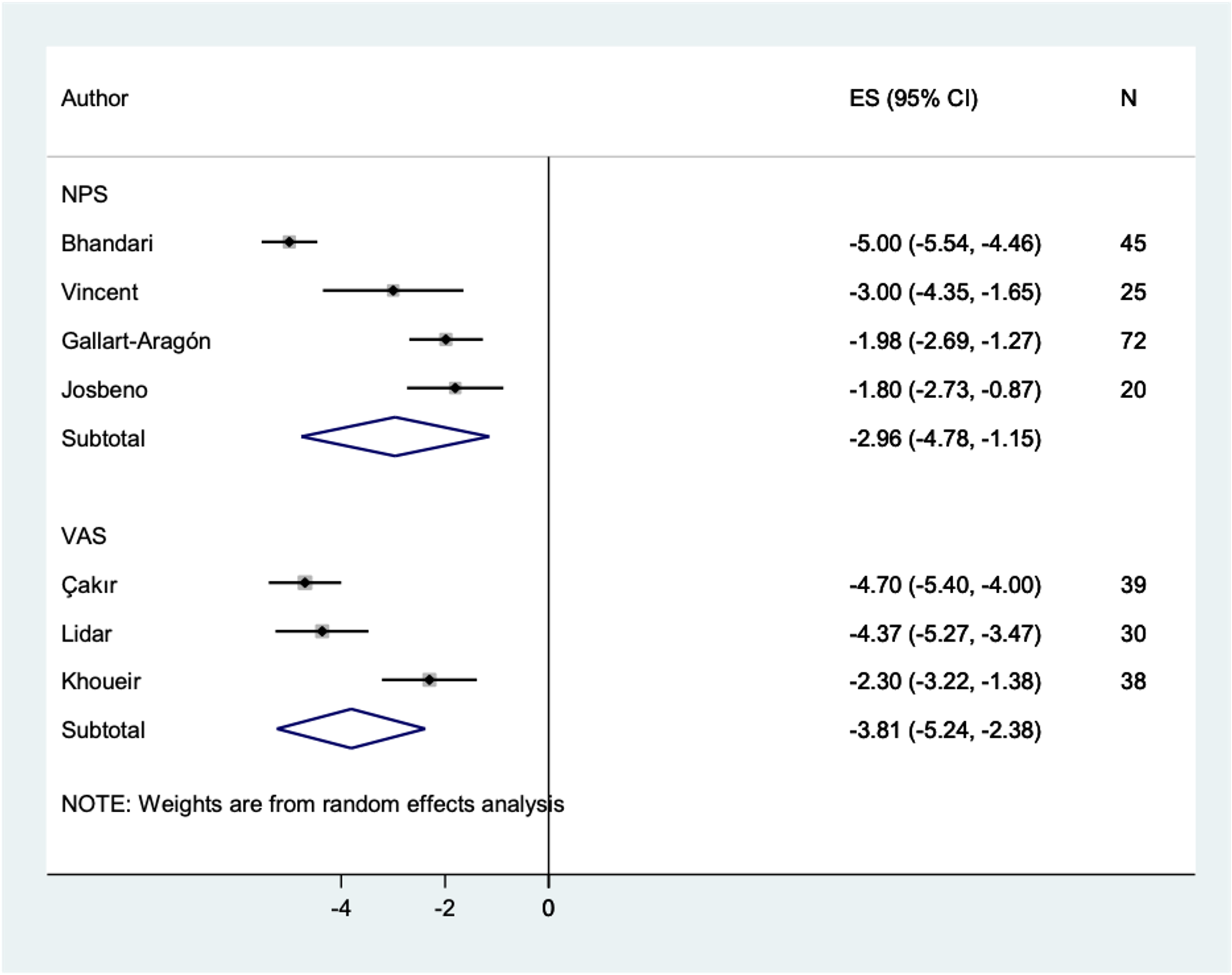
Forest plot of the Random Effect model of pain score changes following bariatric surgery

Four studies measured pain intensity using VAS, ^39, 41, 44^ however one study ^45^ divided the VAS scale in 3 groups: (1) immediately, (2) at its worst, (3) at its best pattern. Therefore, this study was not included in the meta-analysis. Among all patients (with or without back pain) in the three studies, back pain intensity was 3.8 cm lower postoperatively compared to preoperatively. In our meta-analysis the change in VAS-score after bariatric surgery showed a significant change (ES −3.81, 95%CI [−5.24, −2.38], p=0.000) (Fig. 2).

### Subgroup analysis and sensitivity analysis

As there was significant heterogeneity *(I^2^* = 95.1%) of NPS, we obtained subgroup analysis based on publication date, number of patients and follow-up period. It showed no effect on the heterogeneity. A sensitivity analysis of the results showed the heterogeneity was caused by Bhandari *et al.^38^* A re-analysis of the NPS data, resulted in a new forest plot, *I^2^* = 9.2% (Supplemental Digital Content 3_Figure 2 to Supplemental Digital Content 3_Figure 5).

### Random effect model (fixed effect estimate)

As there was significant heterogeneity, but a small included study amount, we obtained the fixed effect estimate for the aggregated mean. Both methods showed significant decreases in pain score.

In our meta-analysis using the fixed estimate, the changes in NPS-score and VAS-score after bariatric surgery showed a significant change (NPS: (ES −3.49, 95%CI [−3.86, −3.12], p=0.001), VAS: (ES −3.98, 95%CI [−4.45, −3.50], p=0.000)) (Fig. 3).

**Fig. 3.**
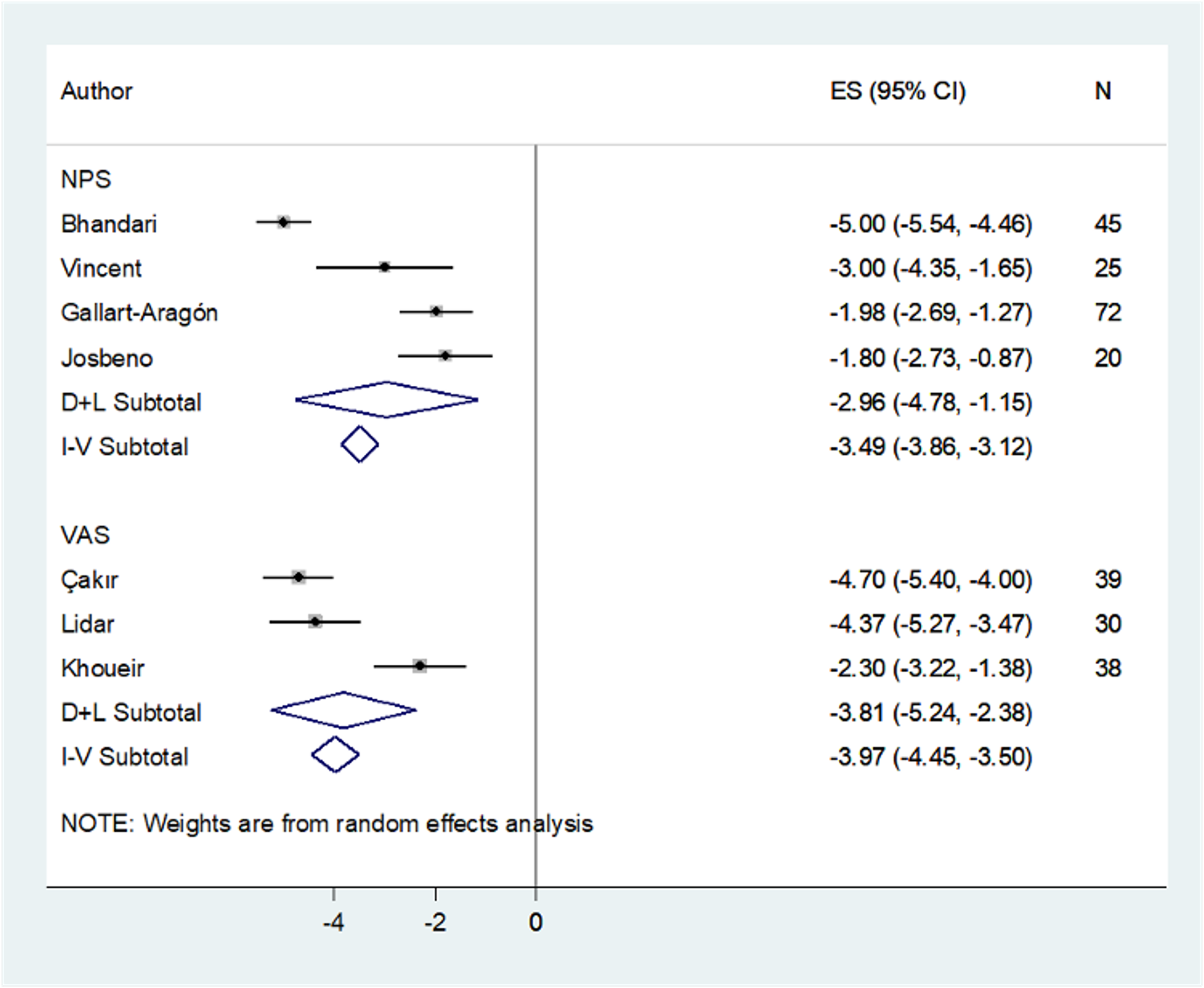
Forest plot random effect model with fixed effect estimate of pain score changes following bariatric surgery.

**Fig. 4.**
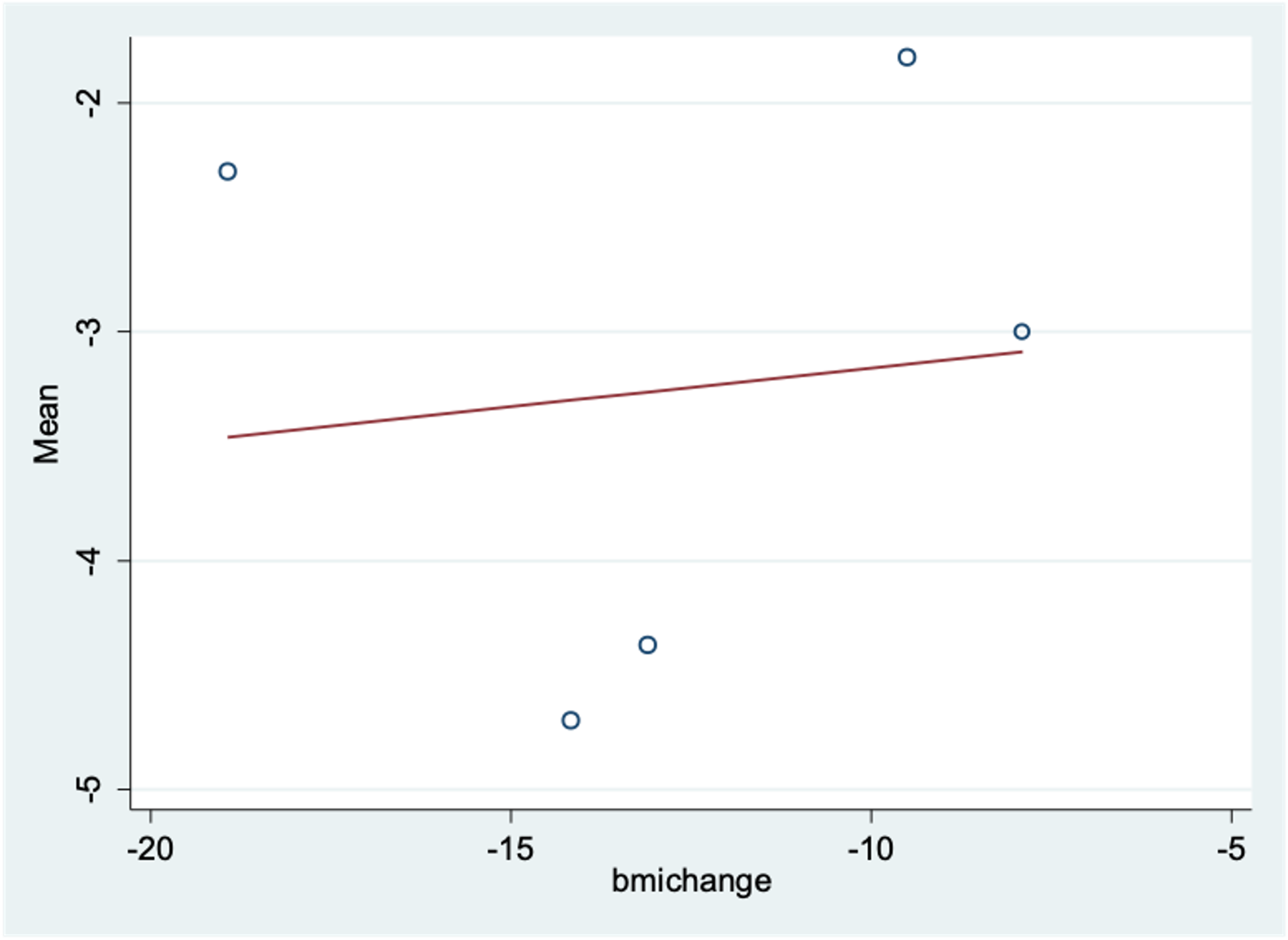
Association of mean pain change with BMI change (X-axis: BMI-change; Y-axis: mean pain change). No significant correlation between BMI and pain changes, r = 0.034.

### Pain intensity score changes compared with a non-surgical control group

One controlled prospective study of 45 morbidly obese patients wanted to examine if patients (n = 25) undergoing laparoscopic Roux-en-Y gastric bypass or laparoscopic adjustable gastric banding demonstrated changes in joint pain, gait, mobility and quality of life after 3 months compared with a nonsurgical group (n = 20).^40^ The researchers found a 16.8% BMI change in the bariatric group, no changes in the control group. The NPS score within the surgery group decreased significantly with 54%, with no change within the control group. The NPS change between the surgical and control-group was significant as well. Furthermore, gait parameters, walking speed and quality of life significantly improved by 3 months after bariatric surgery.

### The association of mean pain change with BMI change

In a total of 318 patients the mean initial BMI was 47.4 kgm^2^ and the reduction in BMI ranged from 16.8% to 30.5%. As shown in Table 1, four studies calculated the correlation between BMI changes and pain intensity improvement. In this meta-analysis to evaluate the association of mean pain change with BMI change, BMI change was included as predictor in a meta-regression. BMI change was available for 5 of the 8 studies. In our meta-analysis no significant relationship between BMI change and decrease in pain score could be established (r = 0.034, 95%CI [−0.52, 0.589], p = 0.19). The results are shown in Figure 4.

### Other back pain outcomes

Five of the eight studies investigated other outcomes for back pain: (a) Rolland-Morris disability questionnaire (RMD), (b) ODI, (c) Waddell disability index, (d) Intervertebral disc space height, and (e) PPT. Most of these measures showed favourable improvement in back pain.

The RMD was examined by two uncontrolled prospective studies. Melissas *et al*. examined 29 obese patients with back pain symptoms undergoing vertical banded gastroplasty with a follow-up of 2 years, to quantify the disability caused by back pain in obese patients and to examine the improvements from weight loss after surgery.^45^ The Rolland-Morris score decreased from 7.9 to 2.0 (p<0.001) postoperatively. Researchers found that after 2-years of follow-up, 34% (n = 10) continued to have back pain symptoms. Bhandari *et al*. examined 45 nonambulatory patients with functional disabilities (walker-dependent, wheelchair-bound, or bedridden) undergoing bariatric surgery to assess functional ability and to determine correlation between BMI, sex, age, comorbidities and functional abilities 12 months postoperatively.^38^. The RMD significantly improved after 12 months, with a p = 0.001.

The ODI was assessed by two uncontrolled studies. Melissas *et al*. showed a decrease in ODI from 21.2 to 5.6 postoperatively (p<0.001).^45^ Khouier *et al*. examined 58 morbidly obese patients with axial back pain (with a history of ≥2 years) undergoing bariatric surgery to examine clinically reported changes in axial low back pain symptoms 12 months after surgery.^39^ The ODI significantly decreased by 24%, scores changed from 26.8 to 20.4 (p = 0.05). The Waddell Disability Index was examined by Melissas *et al*., it decreased significantly from 2.8 to 0.6 postoperatively (p<0.001).

The intervertebral disc space height was examined by Lidar *et al*. in 30 morbidly obese patients undergoing bariatric surgery to determine the effect on axial and radicular pain, intervertebral disc space height, and quality of life one year after surgery.^44^ The disc space hight were assessed from abdominal CT-scans. One year postoperatively the L4-L5 disc space height on CT-scans improved from 6 mm to 8 mm (p<0.001).

The PPT was measured by Gallart-Aragón *et al*. in 72 morbidly obese patients undergoing sleeve gastroplasty to investigate the quality of life and pain. No significant changes were seen in PPTs after SG.^42^

## DISCUSSION

In this meta-analysis, a total of 318 patients that underwent bariatric surgery for the treatment of obesity have been included. The back pain scores were assessed before surgery and during follow-up. All eight included studies reported a reduction of back pain problems following bariatric surgery. Furthermore, substantial weight loss and/or BMI reduction was reported as well. Our meta-analysis showed a significant decrease in pain scores over the eligible studies. However, the heterogeneity in the random effect model by test was significant, 95.1% and 88.7%, indicating that the statistical validity of the summary estimate of effect is affected.

As there was high heterogeneity, subgroup and sensitivity analyses were performed. The sensitivity analyses showed an impact on the heterogeneity by Bhandari and Khoueir *et al*. The different results across the sensitivity analyses, indicate that the result may need to be interpreted with caution. Statistical heterogeneity can be a consequence of clinical or methodological diversity. Clinical diversity is present in every study and is hard to avoid. Among our studies the different surgical weight loss procedures that were used, the diversity in the participants and the diversity in outcomes of the studies were all of influence. The methodological diversity was explained by pain assessment. Pain is a highly subjective measurement experienced individually different by patients. Also, pain assessment differs over time; the score strongly depends on the moment and method of assessment. For example: “the score of the pain now”, or “the highest score experienced in that month” or “that week”, or “the score they experienced during exercise”, are all different. This makes pain difficult to objectively assess and could explain why we see a difference in effect size between the different studies. This methodological diversity could suggests that the studies are not all estimating the same pain intensity, but does not necessarily suggest that the true intervention effect varies ^46^. Therefore, we run a random effect model with fixed effect estimate, this showed a significant decrease in pain scores, with a p<0.05 in both NPS and VAS scores.

When heterogeneity is solely associated with methodological diversity, it indicates that the studies suffer from different degrees of bias. All included studies were prospective cohort studies, because no randomized controlled trials have been published. According to the NOS-tool, seven of the eight studies were at high risk for bias. Furthermore, according to the GRADE-approach, all the included cohorts were defined as very low-quality evidence. However, due to the large magnitude of the effect size, more than 2, the quality was upgraded to low quality (Table 3).

**Table 3.**
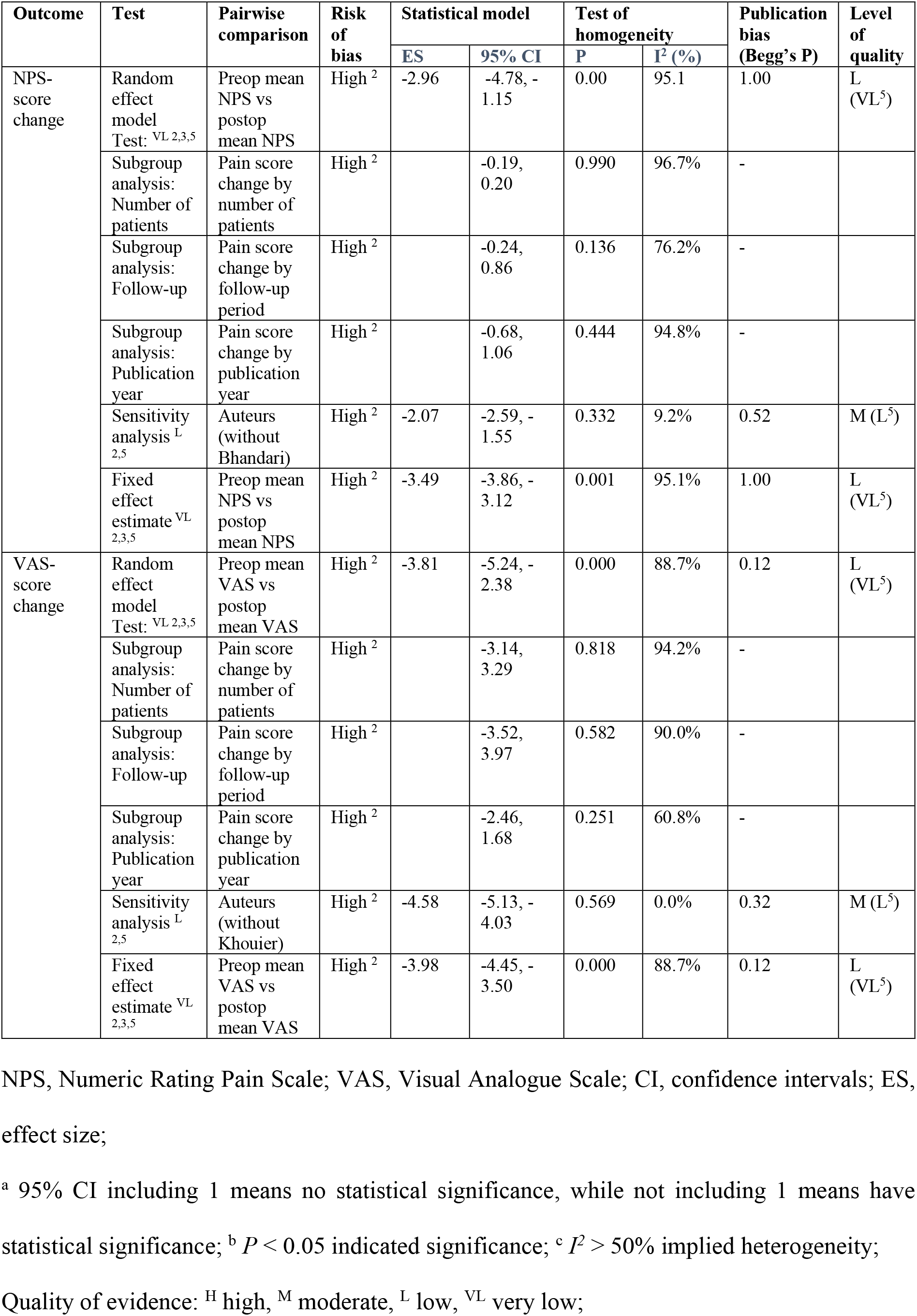
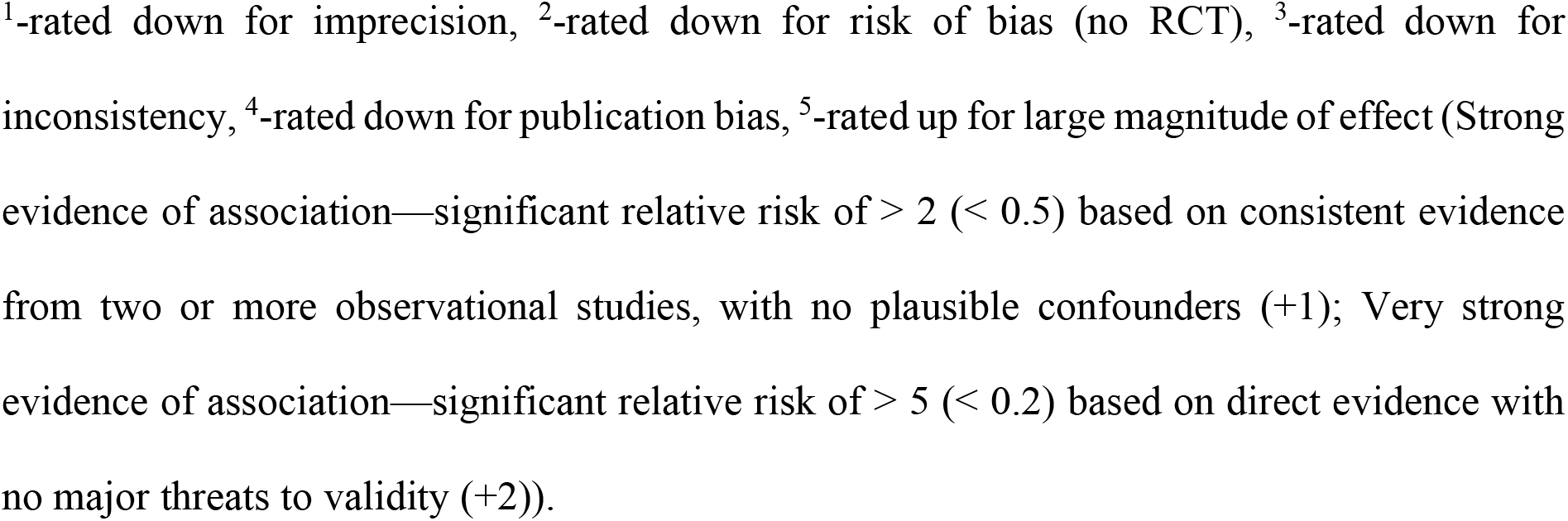
GRADE level of quality assessment

Although one can challenge the validity of these outcomes, reduction of back pain outcomes after surgical weight loss interventions have also been reported by other systematic reviews. A systematic review by Vincent *et al*. looked into back and knee pain improvement after bariatric surgery.^47^ It showed significant reduction of back pain symptoms ranging from 10 - 54%, 1 to 4 years after surgery. Similarly, Joaquim *et al*. showed a significant reduction in back pain symptoms in 10 eligible studies.^48^ Because of the limited number of articles on this subject, there is an overlap between included studies in our meta-analysis. One of the differences with our meta-analysis is that both systematic reviews used multiple back pain assessments as a back pain improvement outcome. To be able to compare the outcomes, we only included NPS and VAS as a primary outcome. Moreover, research has shown that NPS tool is the best tool for neuropathic LBP pain.^49^ To our knowledge this meta-analysis was the first to only take NPS and VAS as an outcome. Next to NPS and VAS, five of the included studies looked into the secondary outcomes and showed favourable results after bariatric surgery. Unfortunately, this data was not sufficient for a meta-analysis. More research is needed to measure back pain relieve more holistically.

Furthermore, as mentioned before a moderate difference in base-line pain scores has been observed. The inclusion criteria among the studies were mostly overlapping, but there was a difference regarding back pain history. Six studies, out of eight, included patients with and without back pain. We observed a higher pain score, if the prevalence of back pain was higher. Unfortunately, three of the six studies failed to report the prevalence of back pain in their study-group, resulting in over- or underestimation of the estimated effect size. The two remaining studies only included obese patients eligible for bariatric surgery with a history of back pain. Both studies did not have a threshold for the pain intensity score for patient inclusion, making overestimation of effect less likely and thus enhancing the value for our analysis. Despite the demonstrated significant pain reduction, the clinical relevance of this reduction remains disputable. Back pain is known to improve in two-third of patients without any interventions.^50^ This makes it difficult to distinguish between reduction of back pain intensity as a result of bariatric surgery or as a result of the natural history of back pain.

Another point of discussion would be the relatively short follow-up period of 3 to 24 months. However, despite this short period, even after 3 months the studies showed a significant decrease in back pain intensity. Studies in obese patients with knee problems have shown similar results. Three months after bariatric surgery, an improvement in knee pain was observed. One of those studies even showed a widening of the knee joint space after 3 months.^51^ The underlying mechanisms of the joint pain improvement are still not completely understood. This early improvement in the back and knee joint, could be an indication for two mechanisms happening: a chronic low-grade inflammation in obese patients and a mechanical compression of the joint. In animal and human models more evidence of a biochemical link between obesity and inflammatory reactions in the joints has been observed.^22^ Another study showed a reduction of inflammatory cytokine levels of interleukin-6 and tumor necrosis factor-α 6 months after weight loss surgery.^32^ However, this study cannot discern between biochemical changes and more research is needed to understand the underlying mechanisms.

Several studies showed a positive association between body weight loss and back pain,^6, 52, 53^ however other studies did not show this association.^39, 44, 54^ Cakir *et al*. showed a significant correlation between pre- and postoperative BMI and rigidity of back pain, with r = 0.98 preoperatively and r = 0.24 postoperatively.^41^ Bhandari *et al*. similarly showed a significant correlation of enhancement of specific functional ability scores and change in BMI, with NPS preoperatively r = −0.29 (p = 0.004) and r = 0.40 (p = 0.002) postoperatively.^38^ Although Koueir *et al*. and Lidar *et al*. found an increasing trend, no significant correlation was established between decrease in BMI and improvement in back pain.^39, 44^ Although not all data in BMI change was available for meta-regression, a meta-regression of five studies showed no significant correlation between BMI change and back pain score changes.

### Limitations

There are several limitations to this study that should be considered. Firstly, this meta-analysis only included prospective observational studies without control groups. The analysis resulted in too much heterogeneity between the pooled studies. A fix effect model showed a significant reduction. However, with the low-quality of the results a definite conclusion cannot be drawn until more studies become available. Next to that, it would have been interesting to compare surgical groups to nonsurgical groups. Only one prospective study included a nonrandomized control group. This nonsurgical group in Vincent *et al*., did not receive the same clinical advice or guidance as the surgical group, making exclusion of other contributing factors more difficult. Secondly, the mean follow-up of the studies was short. No conclusion about long-term effects and the durability of the back pain problems can be made. Thirdly, the history of the patients back pain problems is unknown in most studies. Three of the studies failed to report the prevalence of pre-existing back pain. For most of the studies back pain was not an inclusion criterium and the moment of assessment of back pain in most studies is undescribed. Also, not all included studies reported the loss in patients follow up.

Lastly, as spine surgery, bariatric surgery is highly invasive. It can result in serious complications, such as anastomotic leaks, bleedings, venous thromboembolic - and ischemic events.^55^ Next to that, long term complications as the dumping syndrome, symptomatic cholelithiasis and malnutrition are relevant.^55, 56^ However, despite the risks, surgical interventions give a better outcome in substantial weight reduction and long term effects and research shows an improved quality of life after bariatric surgery.^57, 58^ Still considering the limitations, more research is needed to establish if bariatric surgery should be considered prior to spinal surgery in morbidly obese patients with solely back pain.

## CONCLUSION

From this meta-analysis, the data of back pain improvement following bariatric surgery are encouraging. Substantial weight loss following bariatric surgery might be associated with a reduction in back pain intensity. However, considering the high heterogeneity the evidence is of low-quality. More research is needed to draw a correct weighted conclusion. Ideally a prospective study including spinal imaging, inflammatory markers, longer follow-up period, larger study groups with a randomized control group needs to be done. The relationship between weight loss and reduction in back pain is complex but remains scientifically unclear. To further understand how and why bariatric surgery could reduce the back pain problems of the obese patients, the mechanical and inflammatory effects of obesity on the spine should be better understood.

## Data Availability

Not applicable

## ACKNOWLEDGEMENTS

The authors would like to thank Dr Nancy Briggs (Stats Central, Mark Wainwright Analytical Centre, UNSW) for her help with developing the database search strategy and analysing the data.

---

**Supplemental Digital Content 1.** Search terms used in databases

| Database | Bariatric surgery terms |  | Back pain terms | Search strategy | Results |
| --- | --- | --- | --- | --- | --- |
| Pubmed | (((("Gastric Bypass"[Mesh]) OR ("Gastrectomy"[Mesh]) OR ("Bariatrics"[Mesh]) OR ("Biliopancreatic Diversion"[Mesh]) OR ("Gastroplasty"[Mesh]) OR ("Jejunioleal Bypass"[Mesh]) OR ("Bariatric Surgery"[Mesh]) OR "bariatric surgery" OR "bariatric surgeries" OR "roux-en-y gastric bypass" OR "greenville gastric bypass" OR "gastroileal bypass" OR gastrojejunostomy OR gastrojejunostomies OR gastroplasties OR collis gastroplasty OR vertical-banded gastroplasty OR "vertical banded gastroplasty" OR "vertical banded gastro plas" OR "jejunoileal bypass" OR "ileojejunal bypasses" OR "intestinal bypass" OR "intestinal bypasses" OR "biliopancreatic bypass" OR "biliopancreatic diversion" OR "duodenal switch" OR "pancreatobiliary bypass" OR "gastric banding" OR "stomach banding" OR "swedish gastric banding" OR "swedish adjustable gastric banding" OR "laparoscopic adjustable gastric banding" OR "laparoscopic adjustable silicone banding" OR "bariatric operation" OR "bariatric operations" OR "bariatric procedure" OR "bariatric procedures" OR "bariatric surgical procedure" OR "bariatric surgical procedures" OR "obesity surgery" OR "sleeve gastrectomy" OR "gastric sleeve" OR "lrygb" OR "laparoscopic sleeve gastrectomy" OR SG OR LSG OR "bariatrics" OR "bariatric" OR "metabolic surgery" OR "weight loss surgery" OR GB OR "biliopancreatic diversion" OR "duodenal switch" OR "omega switch" OR "sleeve resection" OR "duodenojejunal bypass" OR "jejunoileal bypass" OR "jejunoileal bypasses" OR "ileojejunal bypass")))) | AND | ((back pain[MeSH terms]) or low back pain[MeSH terms] or back pain* or lumb* pain or lumbago or backache* or back ache* OR "backpain" OR "back pain" OR "Back ache" OR backache OR "lower back pain" OR "Low-back pain" OR "Low back pain" OR "back disorder" OR lumbago OR "lumbar pain" OR Sciatica OR Vertebrogenic Pain Syndrom* OR "musculoskeletal pain")) | #1 AND #2<br>Sort by: **Best Match**<br>Filters: **English** | ➔ 319 articles<br>➔ Title screening: 39 articles<br>➔ Abstract screening: 22<br>➔ Full text 8 |
| MEDLINE | Bariatrics/ OR Bariatric Surgery/ OR Biliopancreatic Diversion/ OR Gastrectomy/ OR Gastric Bypass/ OR Gastroplasty/ OR Jejunioleal Bypass/ OR Anastomosis, Roux-en-Y/ | AND | Back Pain/ OR<br>Low Back Pain/ OR Intervertebral Disc Displacement/ OR Sciatica/ OR<br>Spinal Diseases/ OR<br>Musculoskeletal Pain/ | Filter: English | ➔ 39 studies<br>➔ 7 fitting, 5 maybe (all double)<br>➔ 6 abstract screening<br>➔ 3 fitting full text (double) |
| EMBASE | Bariatric surgery OR bariatric*.mp. OR stomach bypass OR Roux Y anastomosis OR Roux-en-Y gastric bypass OR gastroplasty OR gastric bypass surgery OR jejunioleal bypass OR intestine bypass OR biliopancreatic bypass OR gastric banding OR sleeve gastrectomy OR gastric sleeve OR laparoscopic sleeve gastrectomy | AND | Low back pain OR back pain*.mp. OR backache OR spine disease OR sciatica OR musculoskeletal pain | Filter: English | ➔ 385 articles in total<br>➔ 13 abstracts<br>➔ 8 eligible full text screening (double) |
| CRCC |  |  |  |  |  |
| Other resources (Google scholar and references) | - | - | - | - | - |

**Supplemental Digital Content 2.** Grading of Recommendations Assessment, Development and Evaluation (GRADE) approach for rating the quality of estimates of treatment effect

| GRADE Assessment |
| --- |
| **Ratings**<br>**High quality** ($\oplus\oplus\oplus\oplus$)—We are very confident that the true effect lies close to that of the estimate of the effect<br>**Moderate quality** ($\oplus\oplus\oplus\bigcirc$)— We are moderately confident in the effect estimate: The true effect is likely to be close to the estimate of the effect, but there is a possibility that it is substantially different<br>**Low quality** ($\oplus\oplus\bigcirc\bigcirc$)— Our confidence in the effect estimates is limited: The true effect may be substantially different from the estimate of the effect<br>**Very low quality** ($\oplus\bigcirc\bigcirc\bigcirc$)— We have very little confidence in the effect estimate: The true effect is likely to be substantially different from the estimate of effect |
| **Down rating**<br>The quality rating may be rated down by $-1$ (serious concern) or $-2$ (very serious concern) for the following reasons<br>• Risk of bias (such as failure to conceal random allocation or blind participants in randomised controlled trials or failure to adequately control for confounding in observational studies)<br>• Inconsistency (such as heterogeneity of estimates of effects across trials)<br>• Indirectness (such as surrogate outcomes, study populations or interventions that differ from those of interest, or intransitivity)<br>• Imprecision (for example, 95% confidence intervals are wide and include or are close to null effect)<br>• Publication bias |
| **Up rating**<br>Rating up is typically applied only to observational studies; the most common reason is for a large or very large effect seen over a short period of time and altering a clear downward trajectory |

**Supplemental Digital Content 3_Figure 1.**
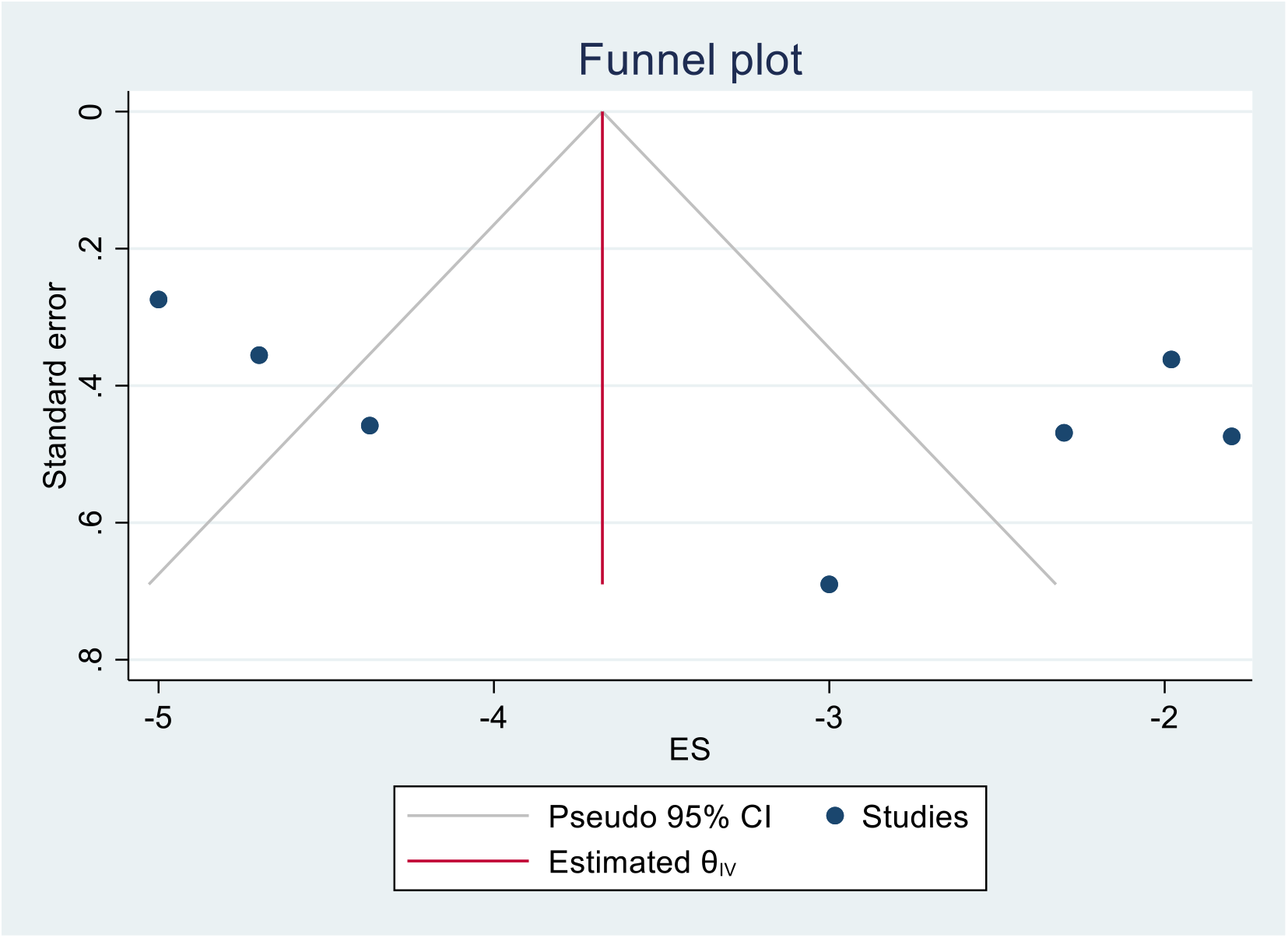
Funnel plot for publication bias (Inverse-Variance method)

**Supplemental Digital Content 3_Figure 2.**
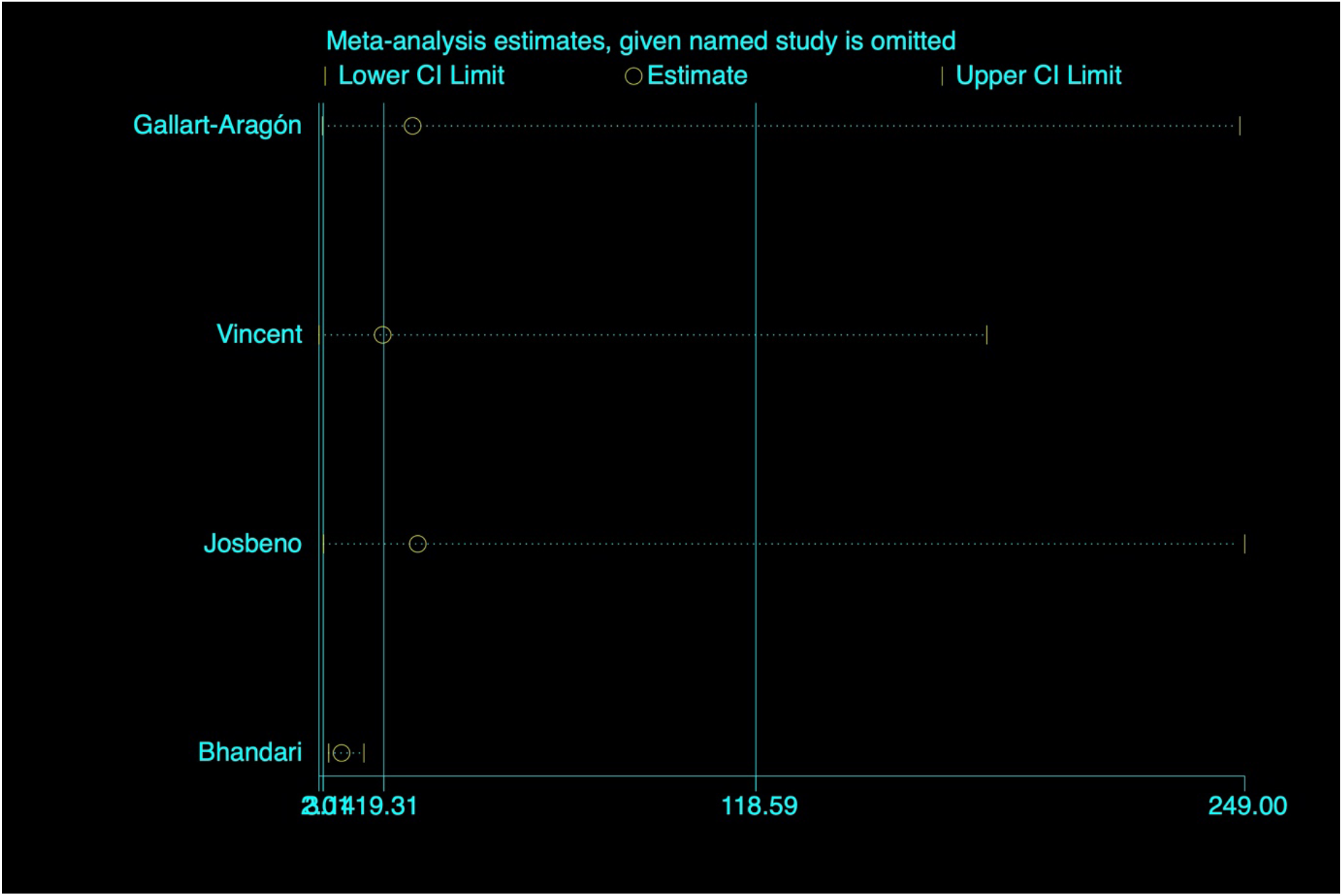
Sensitivity analysis NPS studies.

**Supplemental Digital Content 3_Figure 3.**
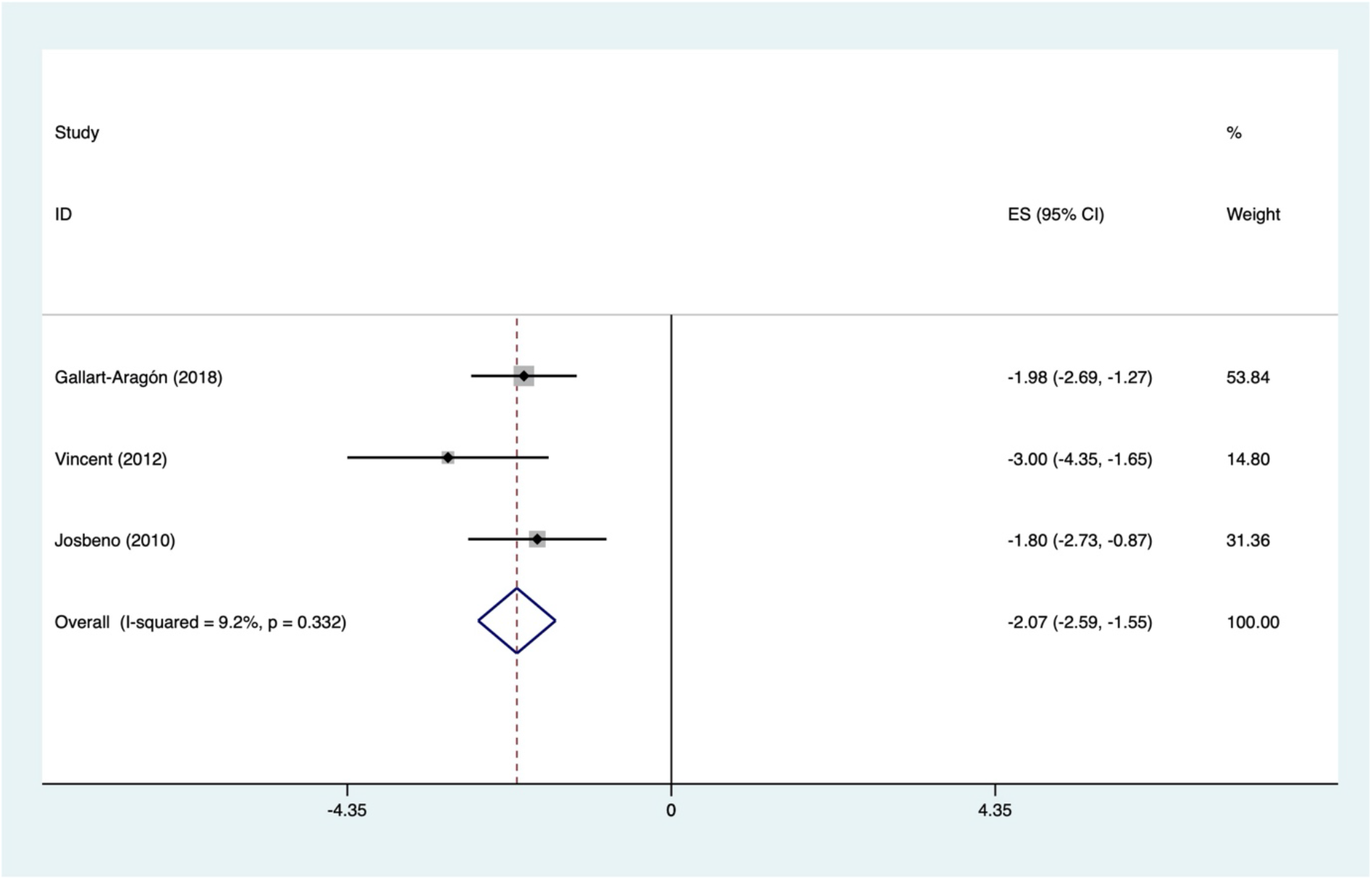
Forest plot NPS score changes after bariatric surgery without Bhandari *et al.*

**Supplemental Digital Content 3_Figure 4.**
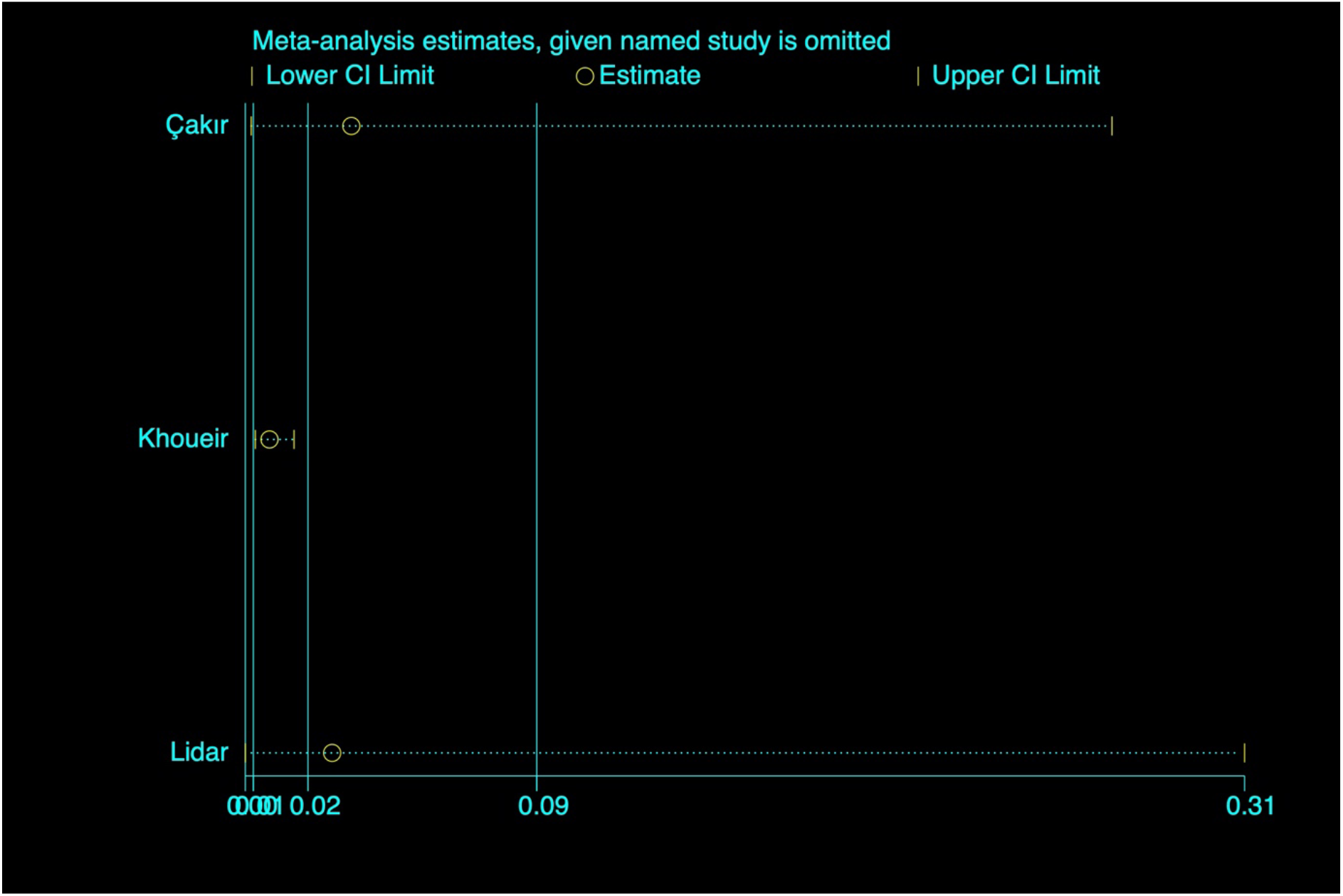
Sensitivity analysis VAS-studies.

**Supplemental Digital Content 3_Figure 5.**
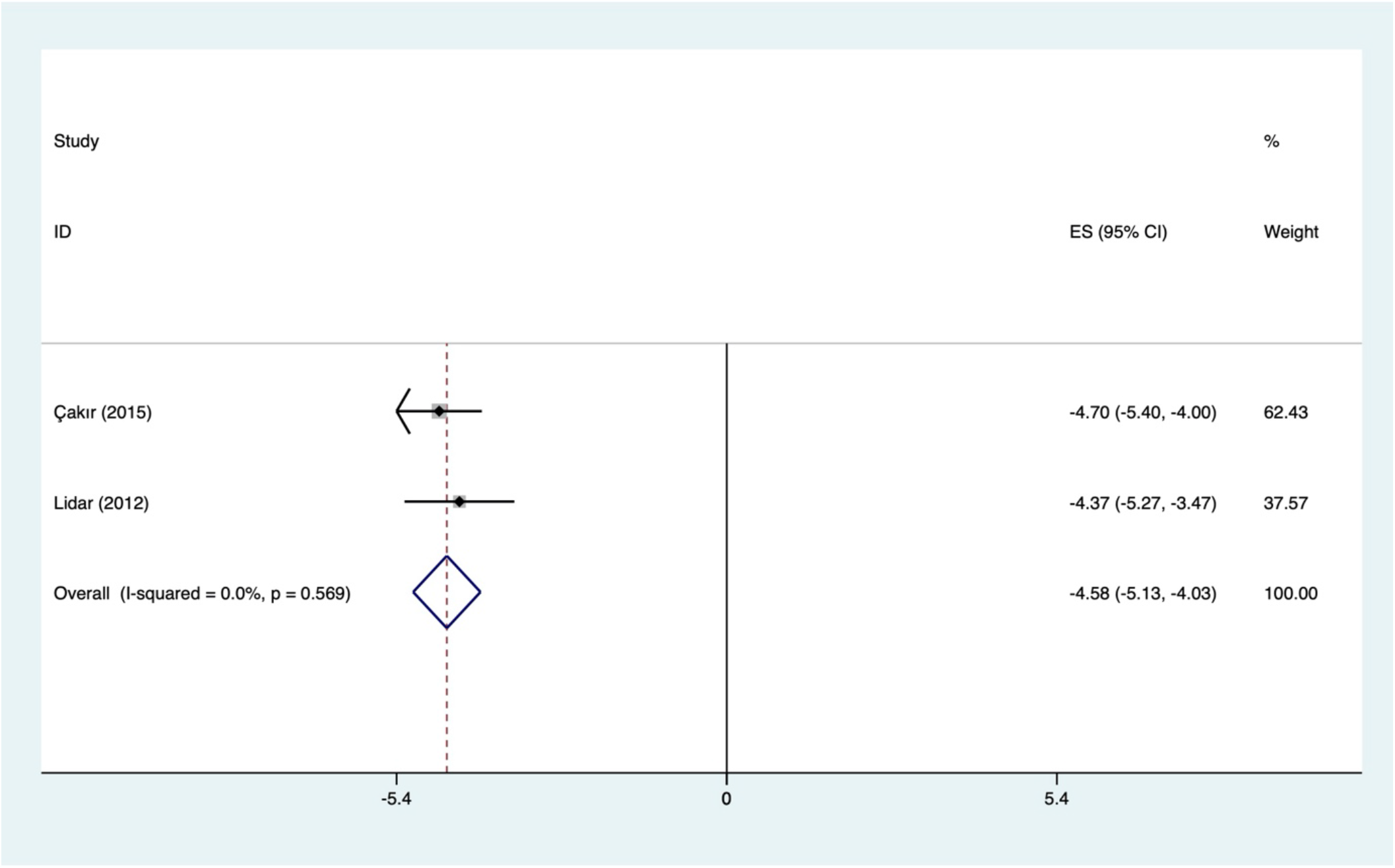
Forest plot VAS score changes after bariatric surgery without Khoueir et al.

## REFERENCES

1. Organization, W. H., Obesity and overweight: Fact sheet. WHO Media Cent. 2016.

2. Goodwin, P. J.; Stambolic, V., Impact of the obesity epidemic on cancer. Annu Rev Med 2015, 66, 281–96.

3. Guh, D. P.; Zhang, W.; Bansback, N.; Amarsi, Z.; Birmingham, C. L.; Anis, A. H., The incidence of co-morbidities related to obesity and overweight: a systematic review and meta-analysis. BMC Public Health 2009, 9, 88.

4. Haslam, D. W. J., W. P. T., Obesity. The Lancet 2005, 366 (9492), 1197–1209.

5. Atchison, J. W.; Vincent, H. K., Obesity and low back pain: relationships and treatment. Pain Manag 2012, 2 (1), 79–86.

6. Shiri, R.; Karppinen, J.; Leino-Arjas, P.; Solovieva, S.; Viikari-Juntura, E., The association between obesity and low back pain: a meta-analysis. Am J Epidemiol 2010, 171 (2), 135–54.

7. Balague, F. M., A. F.; Pellise, F.; Cedraschi, C., Non-specific low back pain. The Lancet 2012, 379 (9814), 482–491.

8. Rubin, D. I., Epidemiology and risk factors for spine pain. Neurol Clin 2007, 25 (2), 353–71.

9. de Schepper, E. I.; Damen, J.; van Meurs, J. B.; Ginai, A. Z.; Popham, M.; Hofman, A.; Koes, B. W.; Bierma-Zeinstra, S. M., The association between lumbar disc degeneration and low back pain: the influence of age, gender, and individual radiographic features. Spine (Phila Pa 1976) 2010, 35 (5), 531–6.

10. Katz, J. N., Lumbar disc disorders and low-back pain: socioeconomic factors and consequences. J Bone Joint Surg Am 2006, 88 Suppl 2, 21–4.

11. Maetzel, A.; Li, L., The economic burden of low back pain: a review of studies published between 1996 and 2001. Best Pract Res Clin Rheumatol 2002, 16 (1), 23–30.

12. Hadjipavlou, A. G.; Tzermiadianos, M. N.; Bogduk, N.; Zindrick, M. R., The pathophysiology of disc degeneration: a critical review. J Bone Joint Surg Br 2008, 90 (10), 1261–70.

13. Hoy, D.; Brooks, P.; Blyth, F.; Buchbinder, R., The Epidemiology of low back pain. Best Pract Res Clin Rheumatol 2010, 24 (6), 769–81.

14. Feng, Y.; Egan, B.; Wang, J., Genetic Factors in Intervertebral Disc Degeneration. Genes Dis 2016, 3 (3), 178–185.

15. Liuke, M.; Solovieva, S.; Lamminen, A.; Luoma, K.; Leino-Arjas, P.; Luukkonen, R.; Riihimaki, H., Disc degeneration of the lumbar spine in relation to overweight. Int J Obes (Lond) 2005, 29 (8), 903–8.

16. Samartzis, D.; Karppinen, J.; Chan, D.; Luk, K. D.; Cheung, K. M., The association of lumbar intervertebral disc degeneration on magnetic resonance imaging with body mass index in overweight and obese adults: a population-based study. Arthritis Rheum 2012, 64 (5), 1488–96.

17. Urquhart, D. M.; Berry, P.; Wluka, A. E.; Strauss, B. J.; Wang, Y.; Proietto, J.; Jones, G.; Dixon, J. B.; Cicuttini, F. M., 2011 Young Investigator Award winner: Increased fat mass is associated with high levels of low back pain intensity and disability. Spine (Phila Pa 1976) 2011, 36 (16), 1320–5.

18. Urquhart, D. M.; Kurniadi, I.; Triangto, K.; Wang, Y.; Wluka, A. E.; O’Sullivan, R.; Jones, G.; Cicuttini, F. M., Obesity is associated with reduced disc height in the lumbar spine but not at the lumbosacral junction. Spine (Phila Pa 1976) 2014, 39 (16), E962–6.

19. Heindel, J. J.; Newbold, R.; Schug, T. T., Endocrine disruptors and obesity. Nat Rev Endocrinol 2015, 11 (11), 653–61.

20. Ouchi, N.; Parker, J. L.; Lugus, J. J.; Walsh, K., Adipokines in inflammation and metabolic disease. Nat Rev Immunol 2011, 11 (2), 85–97.

21. Ruiz-Fernandez, C.; Francisco, V.; Pino, J.; Mera, A.; Gonzalez-Gay, M. A.; Gomez, R.; Lago, F.; Gualillo, O., Molecular Relationships among Obesity, Inflammation and Intervertebral Disc Degeneration: Are Adipokines the Common Link? Int J Mol Sci 2019, 20 (8).

22. Segar, A. H.; Fairbank, J. C. T.; Urban, J., Leptin and the intervertebral disc: a biochemical link exists between obesity, intervertebral disc degeneration and low back pain- an in vitro study in a bovine model. Eur Spine J 2019, 28 (2), 214–223.

23. Risbud, M. V.; Shapiro, I. M., Role of cytokines in intervertebral disc degeneration: pain and disc content. Nat Rev Rheumatol 2014, 10 (1), 44–56.

24. Wuertz, K.; Haglund, L., Inflammatory mediators in intervertebral disk degeneration and discogenic pain. Global Spine J 2013, 3 (3), 175–84.

25. Podichetty, V. K., The aging spine: The role of inflammatory mediators in intervertebral disc degeneration. Cellular and molecular biology 2007, 53 (5), 4–18.

26. Sampara, P.; Banala, R. R.; Vemuri, S. K.; Av, G. R.; Gpv, S., Understanding the molecular biology of intervertebral disc degeneration and potential gene therapy strategies for regeneration: a review. Gene Ther 2018, 25 (2), 67–82.

27. Higgins, D. M.; Mallory, G. W.; Planchard, R. F.; Puffer, R. C.; Ali, M.; Gates, M. J.; Clifton, W. E.; Jacob, J. T.; Curry, T. B.; Kor, D. J.; Fogelson, J. L.; Krauss, W. E.; Clarke, M. J., Understanding the Impact of Obesity on Short-term Outcomes and In-hospital Costs After Instrumented Spinal Fusion. Neurosurgery 2016, 78 (1), 127–32.

28. Olsen, M. A. M., J.; Lauryssen, C.; Polish, L. B.; Jones, M.; Vest, J.; Fraser, V. J., Risk factors for surgical site infection in spinal surgery. Journal of Neurosurgery 2003.

29. Vaidya, R.; Carp, J.; Bartol, S.; Ouellette, N.; Lee, S.; Sethi, A., Lumbar spine fusion in obese and morbidly obese patients. Spine (Phila Pa 1976) 2009, 34 (5), 495–500.

30. Patel, N.; Bagan, B.; Vadera, S.; Maltenfort, M. G.; Deutsch, H.; Vaccaro, A. R.; Harrop, J.; Sharan, A.; Ratliff, J. K., Obesity and spine surgery: relation to perioperative complications. JNeurosurg Spine 2007, 6 (4), 291–7.

31. Arterburn, D.; Wellman, R.; Emiliano, A.; Smith, S. R.; Odegaard, A. O.; Murali, S.; Williams, N.; Coleman, K. J.; Courcoulas, A.; Coley, R. Y.; Anau, J.; Pardee, R.; Toh, S.; Janning, C.; Cook, A.; Sturtevant, J.; Horgan, C.; McTigue, K. M.; Collaborative, P. C. B. S., Comparative Effectiveness and Safety of Bariatric Procedures for Weight Loss: A PCORnet Cohort Study. Ann Intern Med 2018, 169 (11), 741–750.

32. Moschen, A. R.; Molnar, C.; Geiger, S.; Graziadei, I.; Ebenbichler, C. F.; Weiss, H.; Kaser, S.; Kaser, A.; Tilg, H., Anti-inflammatory effects of excessive weight loss: potent suppression of adipose interleukin 6 and tumour necrosis factor alpha expression. Gut 2010, 59 (9), 1259–64.

33. Welbourn, R.; Pournaras, D. J.; Dixon, J.; Higa, K.; Kinsman, R.; Ottosson, J.; Ramos, A.; van Wagensveld, B.; Walton, P.; Weiner, R.; Zundel, N., Bariatric Surgery Worldwide: Baseline Demographic Description and One-Year Outcomes from the Second IFSO Global Registry Report 2013-2015. Obes Surg 2018, 28 (2), 313–322.

34. Moher, D.; Liberati, A.; Tetzlaff, J.; Altman, D. G.; Group, P., Preferred reporting items for systematic reviews and meta-analyses: the PRISMA statement. PLoS Med 2009, 6 (7), e1000097.

35. Higgins, J. P.; Altman, D. G.; Gotzsche, P. C.; Juni, P.; Moher, D.; Oxman, A. D.; Savovic, J.; Schulz, K. F.; Weeks, L.; Sterne, J. A.; Cochrane Bias Methods, G.; Cochrane Statistical Methods, G., The Cochrane Collaboration’s tool for assessing risk of bias in randomised trials. BMJ 2011, 343, d5928.

36. J., W. G. S. B. O. C. D. P., The Newcastle-Ottawa Scale (NOS) for assessing the quality of nonrandomised studies in meta-analyses. Ottawa, ON: Ottawa Hospital Research Institute.

37. Balshem, H.; Helfand, M.; Schunemann, H. J.; Oxman, A. D.; Kunz, R.; Brozek, J.; Vist, G. E.; Falck-Ytter, Y.; Meerpohl, J.; Norris, S.; Guyatt, G. H., GRADE guidelines: 3. Rating the quality of evidence. J Clin Epidemiol 2011, 64 (4), 401–6.

38. Bhandari, M.; Mathur, W.; Kosta, S.; Salvi, P.; Fobi, M., Assessment of functional ability of nonambulatory patients with obesity: after and before bariatric surgery. Surg Obes Relat Dis 2019, 15 (12), 2087–2093.

39. Khoueir, P.; Black, M. H.; Crookes, P. F.; Kaufman, H. S.; Katkhouda, N.; Wang, M. Y., Prospective assessment of axial back pain symptoms before and after bariatric weight reduction surgery. Spine J 2009, 9 (6), 454–63.

40. Vincent, H. K.; Ben-David, K.; Conrad, B. P.; Lamb, K. M.; Seay, A. N.; Vincent, K. R., Rapid changes in gait, musculoskeletal pain, and quality of life after bariatric surgery. Surg Obes Relat Dis 2012, 8 (3), 346–54.

41. Cakir, T.; Oruc, M. T.; Aslaner, A.; Duygun, F.; Yardimci, E. C.; Mayir, B.; Bulbuller, N., The effects of laparoscopic sleeve gastrectomy on head, neck, shoulder, low back and knee pain of female patients. Int J Clin Exp Med 2015, 8 (2), 2668–73.

42. Gallart-Aragon, T.; Fernandez-Lao, C.; Galiano-Castillo, N.; Cantarero-Villanueva, I.; Lozano-Lozano, M.; Arroyo-Morales, M., Improvements in Health-Related Quality of Life and Pain: A Cohort Study in Obese Patients After Laparoscopic Sleeve Gastrectomy. J Laparoendosc Adv Surg Tech A 2018, 28 (1), 53–57.

43. Josbeno, D. A.; Jakicic, J. M.; Hergenroeder, A.; Eid, G. M., Physical activity and physical function changes in obese individuals after gastric bypass surgery. Surg Obes Relat Dis 2010, 6 (4), 361–6.

44. Lidar, Z.; Behrbalk, E.; Regev, G. J.; Salame, K.; Keynan, O.; Schweiger, C.; Appelbaum, L.; Levy, Y.; Keidar, A., Intervertebral disc height changes after weight reduction in morbidly obese patients and its effect on quality of life and radicular and low back pain. Spine (Phila Pa 1976) 2012, 37 (23), 1947–52.

45. Melissas, J.; Kontakis, G.; Volakakis, E.; Tsepetis, T.; Alegakis, A.; Hadjipavlou, A., The effect of surgical weight reduction on functional status in morbidly obese patients with low back pain. Obes Surg 2005, 15 (3), 378–81.

46. Higgins, J. P. T. T. J. C. J. C. M., Li T.; Page M. J. W. V., Cochrane Handbook for Systematic Reviews of Interventions version 6.0 (updated July 2019). Cochrane, 2019. Handbook.

47. Vincent, H. K.; Ben-David, K.; Cendan, J.; Vincent, K. R.; Lamb, K. M.; Stevenson, A., Effects of bariatric surgery on joint pain: a review of emerging evidence. Surg Obes Relat Dis 2010, 6 (4), 451–60.

48. Joaquim, A. F.; Helvie, P.; Patel, A. A., Bariatric Surgery and Low Back Pain: A Systematic Literature Review. Global Spine J 2020, 10 (1), 102–110.

49. Garg, A.; Pathak, H.; Churyukanov, M. V.; Uppin, R. B.; Slobodin, T. M., Low back pain: critical assessment of various scales. Eur Spine J 2020, 29 (3), 503–518.

50. Smith, S. E.; Darden, B. V.; Rhyne, A. L.; Wood, K. E., Outcome of unoperated discogram-positive low back pain. Spine (Phila Pa 1976) 1995, 20 (18), 1997–2000; discussion 2000-1.

51. Abu-Abeid, S.; Wishnitzer, N.; Szold, A.; Liebergall, M.; Manor, O., The influence of surgically-induced weight loss on the knee joint. Obes Surg 2005, 15 (10), 1437–42.

52. Bener, A.; Alwash, R.; Gaber, T.; Lovasz, G., Obesity and low back pain. Coll Antropol 2003, 27 (1), 95–104.

53. Hooper, M. M.; Stellato, T. A.; Hallowell, P. T.; Seitz, B. A.; Moskowitz, R. W., Musculoskeletal findings in obese subjects before and after weight loss following bariatric surgery. Int J Obes (Lond) 2007, 31 (1), 114–20.

54. Mortimer, M.; Wiktorin, C.; Pernol, G.; Svensson, H.; Vingard, E.; Center, M. U-N. s. g. M. I., Sports activities, body weight and smoking in relation to low-back pain: a population-based case-referent study. Scand J Med Sci Sports 2001, 11 (3), 178–84.

55. Ma, I. T.; Madura, J. A., 2nd, Gastrointestinal Complications After Bariatric Surgery. Gastroenterol Hepatol (N Y) 2015, 11 (8), 526–35.

56. Elrazek, A. E.; Elbanna, A. E.; Bilasy, S. E., Medical management of patients after bariatric surgery: Principles and guidelines. World J Gastrointest Surg 2014, 6 (11), 220–8.

57. Major, P.; Matlok, M.; Pedziwiatr, M.; Migaczewski, M.; Budzynski, P.; Stanek, M.; Kisielewski, M.; Natkaniec, M.; Budzynski, A., Quality of Life After Bariatric Surgery. Obes Surg 2015, 25 (9), 1703–10.

58. Mazer, L. M.; Azagury, D. E.; Morton, J. M., Quality of Life After Bariatric Surgery. Curr Obes Rep 2017, 6 (2), 204–210.

